# Prevalence, characteristics, and predictors of Long COVID among diagnosed cases of COVID-19

**DOI:** 10.1101/2022.01.04.21268536

**Authors:** M. C. Arjun, Arvind Kumar Singh, Debkumar Pal, Kajal Das, Alekhya Gajjala, Mahalingam Venkateshan, Baijayantimala Mishra, Binod Kumar Patro, Prasanta Raghab Mohapatra, Sonu Hangma Subba

**Affiliations:** Department of Community Medicine and Family Medicine, All India Institute of Medical Sciences Bhubaneswar, Odisha, India; College of Nursing, All India Institute of Medical Sciences Bhubaneswar, Odisha, India; Department of Microbiology, All India Institute of Medical Sciences Bhubaneswar, Odisha, India; Department of Pulmonary Medicine and Critical Care, All India Institute of Medical Sciences Bhubaneswar, Odisha, India

## Abstract

**Background:** Long COVID or long-term complication after COVID-19 has the ability to affect health and quality of life. Knowledge about the burden and predictors could aid in their prevention and management. Most of the studies are from high-income countries and focus on severe cases. We did this study to estimate the prevalence and identify the characteristics and predictors of Long COVID among our patients.

**Methodology:** We recruited adult (≥18 years) patients who were diagnosed as Reverse Transcription Polymerase Chain Reaction (RTPCR) confirmed SARS-COV-2 infection and were either hospitalized or tested on outpatient basis. Eligible participants were followed up telephonically after four weeks of diagnosis of SARS-COV-2 infection to collect data on sociodemographic, clinical history, vaccination history, Cycle threshold (Ct) values during diagnosis and other variables. Characteristics of Long COVID were elicited, and multivariable logistic regression was done to find the predictors of Long COVID.

**Results:** We have analyzed 487 individual data with a median follow-up of 44 days (Inter quartile range (IQR): 39,47). Overall, Long COVID was reported by 29.2% (95% Confidence interval (CI): 25.3%,33.4%) participants. Prevalence of Long COVID among patients with mild/moderate disease (n = 415) was 23.4% (95% CI: 19.5%,27.7%) as compared to 62.5% (95% CI: 50.7%,73%) in severe/critical cases(n=72). The most common Long COVID symptom was fatigue (64.8%) followed by cough (32.4%). Statistically significant predictors of Long COVID were - Pre-existing medical conditions (Adjusted Odds ratio (aOR)=2.00, 95% CI: 1.16,3.44), having a more significant number of symptoms during acute phase of COVID-19 disease (aOR=11.24, 95% CI: 4.00,31.51), two doses of COVID-19 vaccination (aOR=2.32, 95% CI: 1.17,4.58), the severity of illness (aOR=5.71, 95% CI: 3.00,10.89) and being admitted to hospital (Odds ratio (OR)=3.89, 95% CI: 2.49,6.08).

**Conclusion:** A considerable proportion of COVID-19 cases reported Long COVID symptoms. More research is needed in Long COVID to objectively assess the symptoms and find the biological and radiological markers.

## INTRODUCTION

COVID-19 was declared a pandemic in March 2020. (1) Globally, 263 million people have been diagnosed, and around 5 million are reported dead due to COVID-19. (2) Health systems worldwide are striving to stop the spread of the SAR-COV-2 virus and prevent death and complication due to COVID-19. Apart from acute illness, dialogues on the chronic effect of COVID-19 gained momentum as medical practitioners worldwide started reporting on post COVID complications even in mild cases. (3) It was observed that symptoms of COVID-19 either persist or new symptoms arise after a patient has recovered. Multiple nomenclatures began to appear to describe this condition which includes Long COVID, chronic COVID syndrome, long hauler COVID, post-acute sequelae of COVID-19, post-acute COVID19 syndrome etc. (4,5)

Long COVID was discussed widely, and research was initiated to understand this phenomenon. (6) But there was no widely accepted definition for Long COVID, making it difficult to diagnose and treat the condition. (7) The National Institute for Health and Care Excellence (NICE) was among the first to come out with a rapid guideline to define Long COVID. (8) NICE defines Long COVID as signs and symptoms that continue or develop after acute COVID‐19, including both ongoing symptomatic COVID‐19 (from 4 to 12 weeks) and post‐COVID‐19 syndrome (12 weeks or more). (9) Similarly, the Centers for Disease Control and Prevention (CDC) define Long COVID as a Post-COVID condition with a wide range of new, returning, or ongoing health problems people can experience four or more weeks after first being infected with the virus that causes COVID-19. (10) WHO recently published a document defining the Long COVID based on the Delphi method. (11)

Studies have shown that Long COVID can affect almost all systems in the body. (12) The most described in the literature are respiratory disorders, cardiovascular disorders, neurocognitive disorders, mental health disorders, metabolic disorders etc. Symptoms are in multitudes: including fatigue, breathlessness, cough, anxiety, depression, palpitation, chest main, myalgia, cognitive dysfunction (“brain fog”), loss of smell, etc. (12) Newer symptoms are identified and included in the long COVID as evidence emerges. (13) Many initiatives have been launched to estimate the burden and characteristics of long COVID, especially in developed countries. The Office of National Statistics (ONS) in the United Kingdom gives an estimate of the prevalence and risk factors of long COVID using the national Coronavirus (COVID-19) Infection Survey (CIS). (14) COVID Symptom study app is another data source. (15) National Institute of Health in the USA has also launched new initiatives to study Long COVID and is expected to bring out more evidence. (6) Independent researchers are also working on understanding this phenomenon.

In India, significantly less attention has been given to the burden of Long COVID. (16) During this study’s conception, there were no research papers available in peer-reviewed journals that measured the burden of Long COVID in India. As of today, India has more than 34 million cases of COVID-19. (17) This can translate into a huge number of patients suffering from long COVID. Once the active cases come down, the already overstretched health systems can witness another public health crisis in the form of Long COVID. To mitigate this, we should have a clear idea about Long COVID to develop better management strategies. Post COVID care clinics are already functioning in some parts of the country. (18) The Government of India has also published a guideline for the management of post COVID sequelae. (19) But none of the systematic reviews on long COVID has included a study from India, and there is a wide evidence gap on this condition in India. Thus, it is pertinent that we undertake a study to measure the burden, the characteristics, and the predictors of long COVID in India to bring much-needed insight into this condition.

## METHODOLOGY

We estimated the prevalence, characteristics, and predictors of Long COVID by following up a cohort of patients who were Revere Transcription polymerase chain reaction (RTPCR) positive COVID-19 cases. The study was conducted at All India Institute of Medical Sciences (AIIMS) Bhubaneswar, a tertiary care government hospital and research institute. The study population included adult cases (age ≥18 years) of COVID-19 who were diagnosed with RTPCR test from AIIMS Bhubaneswar from April to September. Individuals less than 18 years and pregnant women were excluded.

We accessed the AIIMS Bhubaneswar COVID-19 screening OPD database and records of patients admitted due to COVID-19. The database was cleaned by removing individuals with missing phone numbers, patients who expired, and those less than 18 years. As per the operational definition based on NICE guidelines, these individuals were contacted through telephone after four weeks from the date of their COVID-19 diagnosis. After taking verbal consent, a detailed telephonic interview was conducted to record the socio-demographic details, past medical history including chronic disease and substance use, acute manifestations of COVID-19, and the treatment received. Participants self-reported their height and weight, and Body Mass Index (BMI) was derived. BMI was classified based on World Health Organization criteria (W.H.O). (20) Data on COVID-19 vaccination history was also collected. This was followed by self-reported Long COVID symptoms and their characteristics which included fatigue, cough, loss of taste and smell, cognitive dysfunction (Brain fog), etc., and an open-ended question. The interview questions were adapted from the W.H.O Global COVID-19 Clinical Platform Case Report Form (CRF) for Post COVID condition (Post COVID-19 CRF). (21) All the data in this study was collected by the post-graduate student authors. Pre-testing of the questionnaire was done, and supervised calls were made before the beginning of actual data collection. The data collected during telephonic interviews were directly entered into EpiCollect5 app. An individual who could not be contacted after two attempts were excluded. The RTPCR cycle threshold (Ct) values during diagnosis of COVID-19 were retrieved from the hospital database to study its association with Long COVID symptoms.

### Sample size and Statistical analysis

The sample size for the study was calculated separately for mild to moderate cases and severe cases. Based on previous estimates, a 20% prevalence was taken for mild to moderate cases and the required sample size was 400. (14) For severe cases we assumed a 50% prevalence of Long COVID, and the sample size was calculated to be 100. A relative precision of 20% was used in both calculations.

Data was collected using EpiCollect5 and imported into Microsoft Excel for cleaning. The data was analyzed in statistical software R (version 3.6.3). Prevalence of Long COVID was determined by the number of participants who self-reported any of the Long COVID symptoms. The self-reported characteristics of symptoms were also given as proportions. The data were analyzed separately for mild to moderate and severe to critical patients. Logistics regression was used to find the predictive factors of Long COVID. Statistical significance for univariable analysis was set at p-value less than 0.05. Multivariable logistic regression was done to obtain an adjusted odds ratio with a 95% confidence interval. Clinically significant, and variables which gives a p-value less than 0.2 in univariable analysis were added to the multivariable model.

### Ethical issues

The institutional ethics committee (IEC) of AIIMS Bhubaneswar granted ethical approval before starting the study (IEC Number: T/IM-NF/CM&FM/21/37). The study was explained to each individual, and a telephonic verbal consent was taken before starting data collection. The consent process was approved by the Institutional Ethics Committee. After data collection, if the participant was found to have symptoms of Long COVID, they were referred to Long COVID OPD in the department of Pulmonary Medicine, AIIMS Bhubaneswar.

## RESULTS

We listed 698 COVID-19 RTPCR positive cases from April to September 2021, out of which 189 patients could not be contacted. A total of 509 individuals were eligible to be included into the study. Consent was denied by nine participants, and thus a total of 500 interviews were conducted successfully. On the preliminary evaluation of data, thirteen entries had wrong dates and were dropped from the final analysis. A final sample of 487 individuals was analyzed. (Figure 1) The median follow-up of participants was 44 days (IQR=39,47)

**Figure 1:**
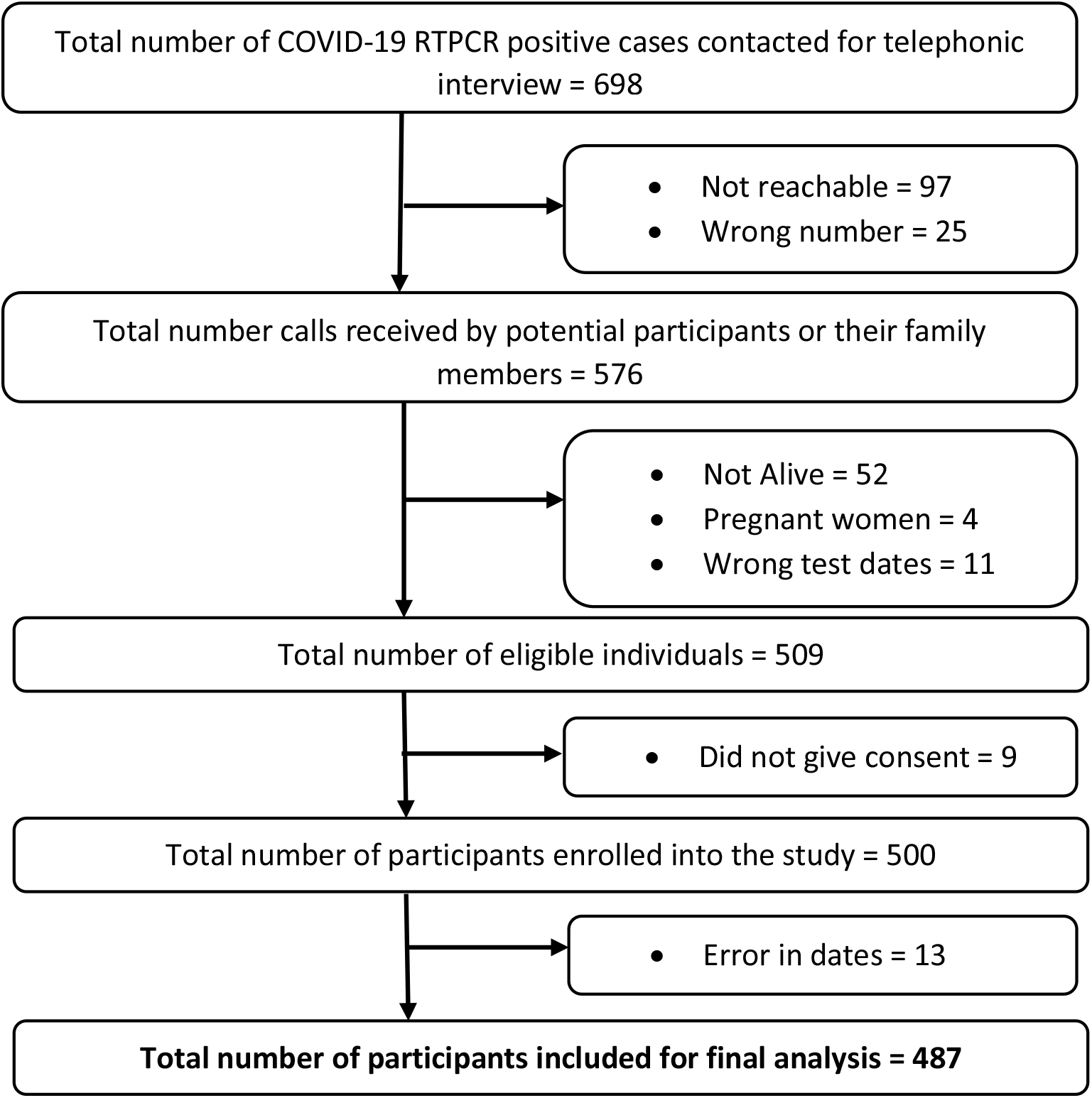
Flow chart showing the selection of study participants

The mean age of the study participants was 39 years (SD=15 years), ranging from 18 to 88 years. Females were 199 (40.9%), and the majority of participants were graduates. Most of the participants were either unemployed or students or homemakers with no earnings. Thirty participants (6.2%) reported being in a job involving COVID-19 management. Majority of participants had normal BMI. (Table 1)

**Table 1:** Sociodemographic characteristics, Past medical history, and vaccination status of participants (n=487)

| **Socio-demographic characteristics** |  |  |
| --- | --- | --- |
| **Variable** |  | **n (%)** |
| Age (Mean (SD); Range) |  | 39 (15); 18 to 88 |
| Females |  | 199 (40.9) |
| Education | Illiterate/No formal education | 24 (4.9) |
|  | Studied up to 10 std or below | 116 (23.8) |
|  | Higher secondary | 75 (15.4) |
|  | Graduate | 218 (44.8) |
|  | Post-graduate and above | 54 (11.1) |
| Current Occupation | Unemployed/Student/Homemaker | 205 (42.1) |
|  | Professionals/Technical/Administrators | 149 (30.6) |
|  | Skilled and Unskilled Manual laborer | 54 (11.1) |
|  | Retired | 11 (2.3) |
|  | Other | 68 (13.9) |
| Occupation involving COVID-19 management |  | 30 (6.2) |
| BMI<br>(n=484) | Underweight (<18.5) | 24 (5) |
|  | Normal (18.5-24.9) | 284 (58.7) |
|  | Overweight (25.0-29.9) | 153 (31.6) |
| | Obese ($\geq 30.0$) | 23 (4.7) |
| **Past Medical History** |  |  |
| History of COVID-19 before the current episode |  | 18 (3.7) |
| Diagnosed to have Diabetes |  | 58 (11.9) |
| Diagnosed to have Hypertension |  | 51 (10.5) |
| Diagnosed to have Anxiety |  | 3 (0.6) |
| Diagnosed to have Asthma |  | 15 (3.1) |
| Diagnosed to have Tuberculosis |  | 5 (1) |
| Diagnosed to have Cancer |  | 19 (3.9) |
| Diagnosed to have other medical condition |  | 53 (10.9) |
| Participants who gave history of substance use |  | 54 (11.1) |
| Smoking status | Current smoker | 16 (3.3) |
|  | Former (Not smoked more than one year) | 7 (1.4) |
| Chewable tobacco |  | 32 (6.6) |
| Alcohol use |  | 19 (3.9) |
| **COVID-19 Vaccination status** |  |  |
| Participants who received two doses of COVID-19 vaccine |  | 287 (58.9) |
| Participants who received one dose of COVID-19 vaccine |  | 81 (16.6) |
| Participants who did not receive COVID-19 vaccine |  | 119 (24.5) |

Eighteen participants (3.7%) reported that they had COVID-19 before the current episode. Around 10% had a history of pre-existing Diabetes or Hypertension. Few participants gave the history of other comorbidities like asthma, tuberculosis, anxiety, cancer, or other chronic diseases, and none reported depression. A single question was used to record any type of self-reported substance use, and 54 (11.1%) participants gave the history of some form of substance use which included alcohol and tobacco. Two doses of vaccine were taken by 287 (58.9%) participants, one dose by 81 (16.6%), and there were 119 (24.5%) who had not been vaccinated at all. The majority of the sample had taken Covaxin. Very few participants reported having side effects post-vaccination.

Clinical features of the participants revealed that majority of them had 1 to 4 symptoms during the acute phase, and the most common symptoms were Fever and Cough. According to W.H.O Post COVID-19 Case Report Form criteria, 415 (85.2%) had mild to moderate and 68 (14%) had severe disease, and four participants (0.8%) had become critical. Most of the participants, 377 (77.4%), underwent home-isolation and were treated as Outpatient and 110 (22.6%) were hospitalized. (Table 2)

**Table 2:**
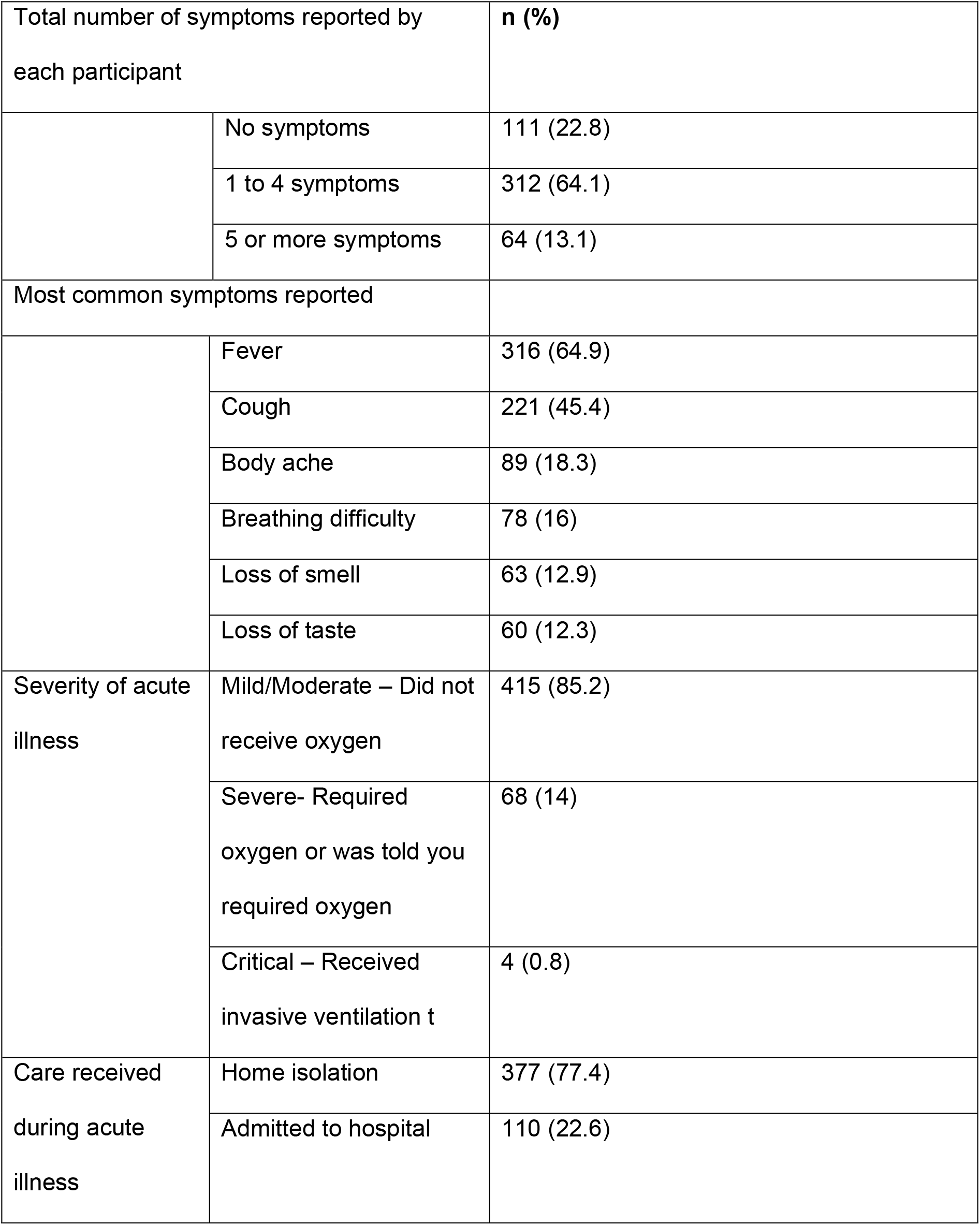
Clinical features and management of acute illness of COVID-19 among participants (n=487)

Overall prevalence of Long COVID was 29.2% (95% CI: 25.3%,33.4%) with 142 individuals reporting it. Subgroup analysis revealed that prevalence of Long COVID was 62.5% (95% CI: 50.7%,73%) among severe/critical cases (n=72), which was significantly higher than among mild/moderate cases (n=415) at 23.4% (95% CI: 19.5%,27.7%). (Figure 2) Among participants who were asymptomatic during the acute phase of COVID-19 (n=111), only six reported Long COVID symptoms.

**Figure 2:**
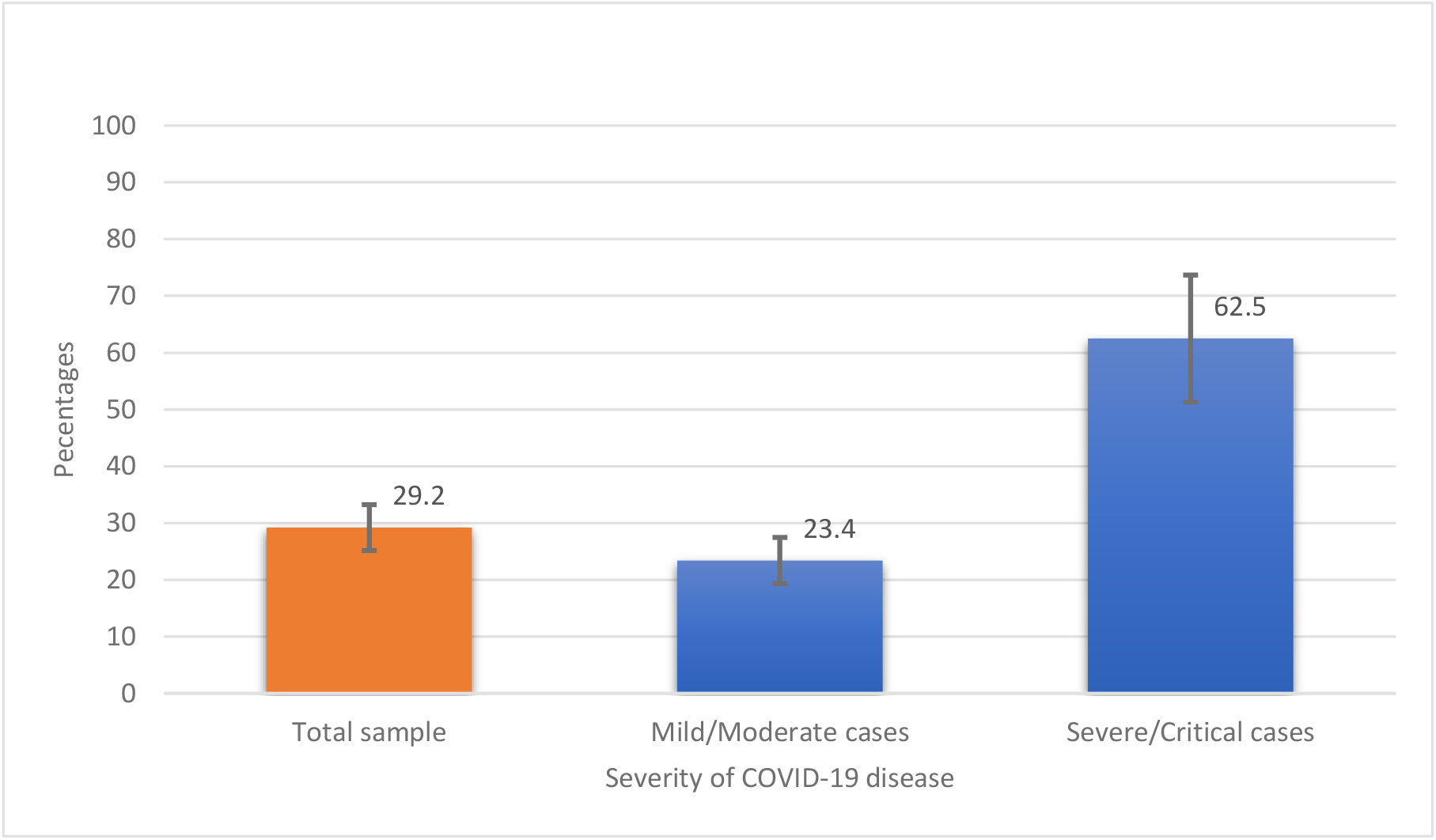
Proportion of self-reported Long COVID symptoms

The most common symptom reported was Fatigue 92 (64.8%), followed by Cough 46 (32.4%). Only three participants reported cognitive dysfunction or Brain fog. Limitation of daily activity following Long COVID was not reported by the majority, but 41 (28.9%) participants reported having some activity limitation. Out of the 142 participants who self-reported Long COVID, 131 (92.3%) perceived the symptoms to be not severe, whereas 11 (7.7%) experienced the symptoms a lot. Health care practitioners were consulted for Long COVID by 49 (34.5%) participants. (Table 3)

**Table 3:**
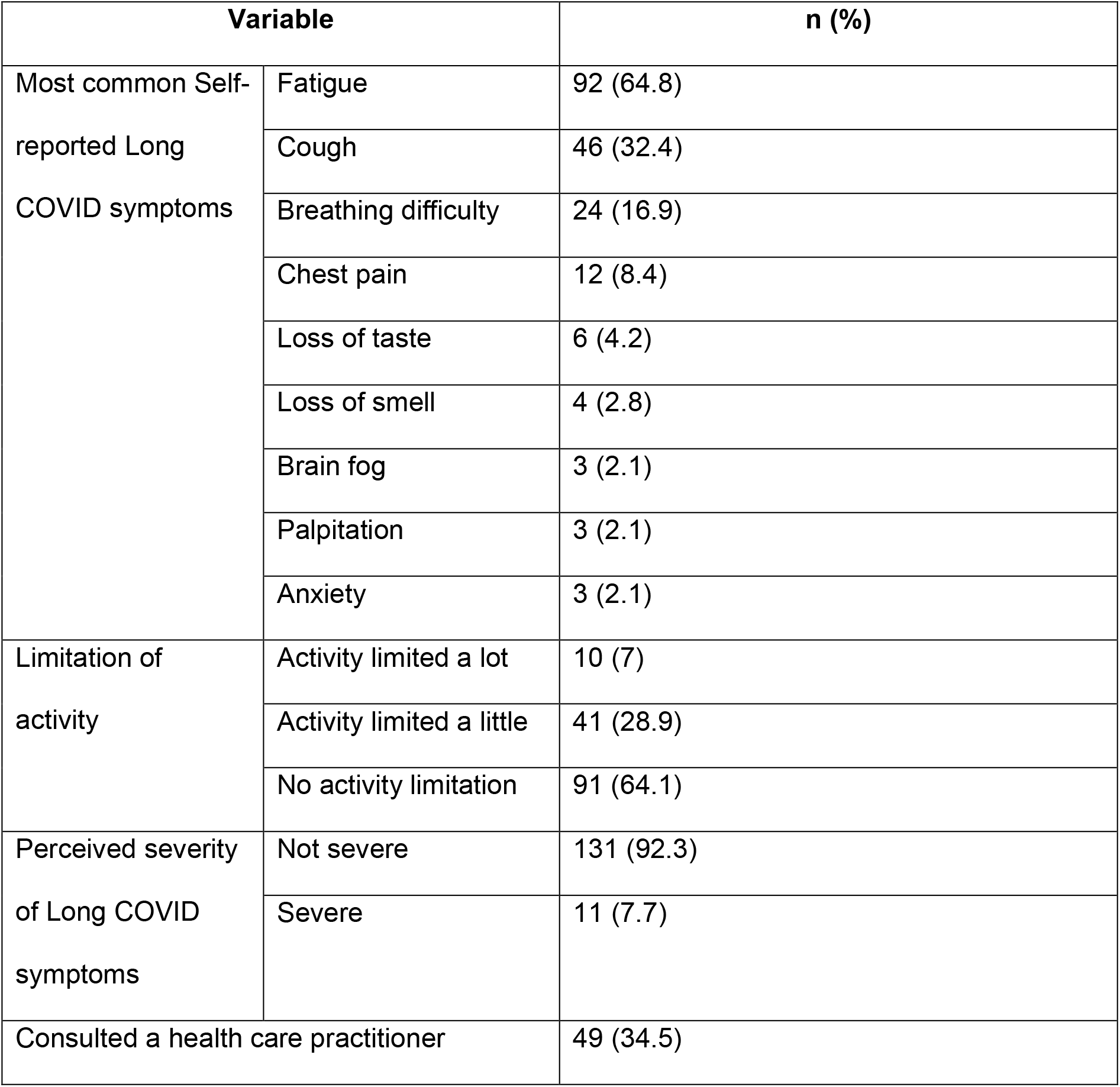
Self-reported Long COVID symptoms and its features (n=142)

| Variable |  | n (%) |
| --- | --- | --- |
| Most common Self-reported Long COVID symptoms | Fatigue | 92 (64.8) |
|  | Cough | 46 (32.4) |
|  | Breathing difficulty | 24 (16.9) |
|  | Chest pain | 12 (8.4) |
|  | Loss of taste | 6 (4.2) |
|  | Loss of smell | 4 (2.8) |
|  | Brain fog | 3 (2.1) |
|  | Palpitation | 3 (2.1) |
|  | Anxiety | 3 (2.1) |
| Limitation of activity | Activity limited a lot | 10 (7) |
|  | Activity limited a little | 41 (28.9) |
|  | No activity limitation | 91 (64.1) |
| Perceived severity of Long COVID symptoms | Not severe | 131 (92.3) |
|  | Severe | 11 (7.7) |
| Consulted a health care practitioner |  | 49 (34.5) |

Analysis for predictors revealed that age, sex, occupation, BMI, history of substance use, and Cycle threshold (Ct) values were not significantly associated with Long COVID. Pre-existing medical conditions (adjusted Odds ratio (aOR) = 2.00 (95% CI: 1.16,3.44)), receiving two doses of COVID-19 vaccination (aOR=2.32 (95% CI: 1.17,4.58)), having more severe COVID-19 disease (aOR = 5.71 (95% CI: 3.00,10.89)) and having a greater number of symptoms during acute phase of COVID-19 disease were significantly associated with Long COVID. Admission to hospital during the acute phase of disease was significantly associated with Long COVID (Odds ratio = 3.89 (95% CI: 2.49,6.08)); however, this variable was not included in the multivariable model due to the colinear relation with the severity of the disease. (Table 4)

**Table 4:** Predictors of Self-reported Long COVID symptoms

| Variable |  | Univariable<br>logistic regression |  | Multivariable<br>logistic regression |  |
| --- | --- | --- | --- | --- | --- |
|  |  | Odds Ratio<br>(95% CI) | p<br>value | Adjusted<br>Odds Ratio<br>(95% CI) | p<br>value |
| Age<br>Categories | 18 to 45 years | Reference | - | Reference | - |
|  | 46 to 59 years | 1.46<br>(0.91,2.36) | 0.12 | 1.24<br>(0.68,2.29) | 0.48 |
|  | 60 years & above | 1.46<br>(0.79,2.67) | 0.22 | 1.08<br>(0.48,2.43) | 0.86 |
| Sex | Male | Reference | - | Reference | - |
|  | Female | 1.33<br>(0.89,1.97) | 0.16 | 1.29<br>(0.74,2.25) | 0.36 |
| Occupation | Unemployed/Student/Homemaker | Reference |  | Reference |  |
|  | Professional/Technical/<br>Administrative/Managerial | 1.32<br>(0.84,2.08) | 0.23 | 1.79<br>(0.96,3.33) | 0.06 |
|  | Skilled/Unskilled manual | 0.65<br>(0.31,1.34) | 0.24 | 0.82<br>(0.32,2.09) | 0.68 |
|  | Other | 0.98<br>(0.55,1.74) | 0.94 | 1.15<br>(0.53,2.48) | 0.73 |
| BMI | Underweight (<18.5) | Reference | - | Reference | - |
|  | Normal or lean (18.5-24.9) | 2.13<br>(0.71,6.44) | 0.18 | 1.58<br>(0.39,6.47) | 0.52 |
|  | Overweight (25.0-29.9) | 2.35<br>(0.76,7.26) | 0.14 | 1.49<br>(0.35,6.26) | 0.58 |
| | Obese ($\geq 30.0$) | 1.05<br>(0.23,4.82) | 0.95 | 0.56<br>(0.09,3.44) | 0.53 |
| History of substance use |  | 0.75<br>(0.39,1.44) | 0.38 | 0.95<br>(0.41,2.16) | 0.89 |
| Past history of COVID-19 |  | 0.93<br>(0.33,2.66) | 0.90 | 0.66<br>(0.20,2.15) | 0.49 |
| Pre-existing medical condition |  | 1.69<br>(1.12,2.55) | 0.01 | 2.00<br>(1.16,3.44) | **0.01** |
| COVID-19 vaccination | Not vaccinated | Reference | - | Reference | - |
|  | Completed 1 dose | 1.30<br>(0.66,2.55) | 0.45 | 1.88<br>(0.84,4.22) | 0.13 |
|  | Completed 2 doses | 2.05<br>(1.23,3.42) | 0.01 | 2.32<br>(1.17,4.58) | **0.01** |
| Number of COVID-19 symptoms | No symptoms | Reference | - | Reference | - |
|  | 1 to 4 symptoms | 9.40<br>(3.99,22.09) | **<0.001** | 6.88<br>(2.74,17.23) | **<0.001** |
|  | 5 or more symptoms | 12.77<br>(4.89,33.37) | **<0.001** | 11.24<br>(4.00,31.51) | **<0.001** |
| Severity of COVID-19 disease | Mild/Moderate | Reference | - | Reference |  |
|  | Severe/Critical | 5.46<br>(3.22,9.27) | **<0.001** | 5.71<br>(3.00,10.89) | **<0.001** |
| Care received during COVID-19 disease | Home Isolation | Reference | - | - | - |
|  | Admitted to hospital | 3.89<br>(2.49,6.08) | **<0.001** | - | - |
| Cycle threshold | E Gene/N Gene (n=442) | 0.98<br>(0.94,1.02) | 0.38 | - | - |
|  | ORF1a/ORF1b/N/N2 Gene<br>(n=378) | 0.99<br>(0.95,1.03) | 0.53 | - | - |

## DISCUSSION

Long COVID is studied extensively all around the world, but research from India is limited. A recently published living systematic review has identified important research gaps, which includes paucity of evidence from low to middle-income countries and in people who were not hospitalized. (22) Both these research gaps are addressed in our study.

The overall prevalence of Long COVID in our study was 29.2%, with a median follow-up period of 44 days. This is comparable to the Office of National Statistics (UK) estimates based on their National Coronavirus (COVID-19) Infection Survey. The survey estimates that around 1 in 5 respondents testing positive for COVID-19 exhibit symptoms for five weeks or longer, i.e., 21% (CI: 19.9,22.1). (14) In mild to moderate cases, the symptoms of Long COVID were 23.4% in our study, after four weeks of COVID-19 infection. A study from India reported Long COVID symptoms in mild COVID-19 to be 22.6% (prevalence of fatigue), albeit with a low sample size. (23) Similarly, another study from Northern India, which followed up the patients from a tertiary care hospital, estimates that 22% had Long COVID. (24) In severe to critical cases with a sample size of 72, our study estimated a prevalence of 62.5%. These estimates are similar to a study from India published in pre-print server, which reported dyspnea in 74.3% and fatigue and disturbed sleep in more than 50% of patients after 30 to 40 days of recovery. (25) Another study in pre-print, which estimated Long COVID in hospitalized patients of North India, gave an estimate of 40.3% after 4 to 6 weeks follow-up. (26) High prevalence of Long COVID symptoms in severe and hospitalized cases are reported from multiple studies from all over the world. (27,28)

The most common Long COVID symptoms found in our study were fatigue and cough. This is similar to other studies from India. (23,24,25,29) The Self-reported symptoms in the COVID Symptom Study app and the National Coronavirus (COVID-19) Infection Survey (CIS) from the United Kingdom has also recorded that fatigue is the most common symptom reported. (14,15) Multiple systematic reviews and meta-analysis on Long COVID have listed fatigue as the most common or among the first three Long COVID symptoms. (12,22,30,31,32) A recent study from India reported fatigue to be present even after three months of recovery from COVID-19. (33) Although fatigue was self-reported in this study, a consistent finding in multiple studies indicates that fatigue is, in fact, the most common of Long COVID symptoms. (34)

Predictors of Long COVID are important because it helps to prioritize the at-risk population and design interventions. In our study, one of the strongest predictors of Long COVID was the severity of COVID-19 disease and hospital admission. This is intuitive because the chances of having persistent symptoms after four weeks post-infection can be higher if the disease is severe. This is backed up by a systematic review which found that hospitalization during the acute infection (odds ratio [OR] 2·9, 95% CI 1·3–6·9) was the most significant predictor of developing the post-COVID syndrome. (31) Similar to the severity of the COVID-19 disease, having more than one symptom during the acute phase of COVID-19 disease was associated with Long COVID. This finding is similar to the COVID symptoms app study based on self-reported symptoms. (15) Another important predictor of Long COVID was the presence of pre-existing conditions like diabetes and hypertension. A study from India and a systematic review on this topic has found a similar and strong association between the pre-existing condition and Long COVID. (25,30) An observational paradox in our study was that the participants who took two doses of COVID-19 vaccination had higher odds of developing Long COVID. It could be due to better survival in vaccinated individuals who may continue to exhibit symptoms of COVID-19 disease. But we could not find any literature on this association, and based on this study, we cannot imply causation. Age and sex, which was commonly found to be associated with Long COVID was not a significant predictor in our study. Cycle threshold (Ct) values of two genes were also not a significant predictor of Long COVID.

The strength of this study is that all the cases of COVID-19 were diagnosed with RTPCR, and there is minimal risk of misclassification. The questionnaire used to capture the Long COVID was adapted from the standard case reporting format recommended by W.H.O. The data were collected by doctors involved in patient care and which improves the validity of the findings. Our study also had limitations. The Long COVID symptoms were all self-reported, and thus objective assessment of symptoms like fatigue was not done. Telephonic interviews precluded us from collecting additional information like clinical and radiological examination for correlating with the findings. We assessed Long COVID after four weeks, and no further follow-up was done. The cause of death of fifty-two individuals who were not alive during the time of data collection was not enquired, and their death may have been related to Long covid complications.

In developed countries, many large-scale cohort studies are undertaken to understand this phenomenon. (35,36) Similar studies on Long COVID are lacking in India, and our research community should bridge this gap. We need more research into Long COVID to objectively assess the symptoms, to monitor the symptoms for a longer duration, and to study the biological and radiological markers, which can lead to better treatment guidelines and comprehensive management of COVID-19 disease.

## Data Availability

All data produced in the present study are available upon reasonable request to the authors. The data will be made available online on a later date.

## Notes

### Competing Interest Statement

The authors have declared no competing interest.

### Funding Statement

This study did not receive any funding

### Author Declarations

The Institutional Ethics Committee (IEC) of All India Institute of Medical Sciences (AIIMS) Bhubaneswar granted ethical approval before starting the study (IEC Number: T/IM-NF/CM&FM/21/37).

## REFERENCES

1. WHO Director-General’s opening remarks at the media briefing on COVID-19 - 30 March 2020 [Internet]. 2020 [cited 2021 Dec 13]. Available from: https://www.who.int/director-general/speeches/detail/who-director-general-s-opening-remarks-at-the-media-briefing-on-covid-1930-march-2020.

2. WHO Coronavirus (COVID-19) Dashboard | WHO Coronavirus (COVID-19) Dashboard with Vaccination Data [Internet]. 2021 [cited 2021 Dec 13]. Available from: https://covid19.who.int/.

3. World health organization. The latest on the covid-19 global situation & long-term sequelae [Internet]. 2021 [cited 2021 Dec 13]. Available from: https://www.who.int/publications/m/item/update-54-clinical-long-term-effects-of-covid-19.

4. Callard F, Perego E. How and why patients made Long Covid. Soc Sci Med. 2021;268:113426.

5. Baig AM. Chronic COVID syndrome: Need for an appropriate medical terminology for long-COVID and COVID long-haulers. J Med Virol. 2021;93(5):2555–6.

6. Collins FS. NIH launches new initiative to study “Long COVID” | National Institutes of Health (NIH) [Internet]. 2021 Feb [cited 2021 Dec 13]. Available from: https://www.nih.gov/about-nih/who-we-are/nih-director/statements/nih-launches-new-initiative-study-long-covid.

7. Sifferlin A. How Covid-19 Long Haulers Created a Movement | by Alexandra Sifferlin | Medium Coronavirus Blog. 2020 Nov 11 [cited 2021 Dec 13]; Available from: https://coronavirus.medium.com/how-covid-19-long-haulers-created-a-movement-24313746833a.

8. Venkatesan P. NICE guideline on long COVID. Lancet Respir Med. 2021;9(2):129.

9. Overview | COVID-19 rapid guideline: managing the long-term effects of COVID-19 | Guidance | NICE. 2020 Dec 18 [cited 2021 Dec 13]; Available from: https://www.nice.org.uk/guidance/ng188.

10. Post-COVID Conditions | CDC [Internet]. 2021 Sep [cited 2021 Dec 13]. Available from: https://https://www.cdc.gov/coronavirus/2019-ncov/long-term-effects/index.html.

11. A clinical case definition of post COVID-19 condition by a Delphi consensus, 6 October 2021 [Internet]. 2021 [cited 2021 Dec 13]. Available from: https://www.who.int/publications/i/item/WHO-2019-nCoV-Post_COVID-19_condition-Clinical_case_definition-2021.1.

12. Lopez-Leon S, Wegman-Ostrosky T, Perelman C, Sepulveda R, Rebolledo P, Cuapio A et al. More than 50 long-term effects of COVID-19: a systematic review and meta-analysis. Scientific Reports. 2021;11(1).

13. Raj SR, Arnold AC, Barboi A, Claydon VE, Limberg JK, Lucci VM, et al. Long-COVID postural tachycardia syndrome: an American Autonomic Society statement. Clin Auton Res. 2021;31(3):365–8.

14. Office for National Statistics. The prevalence of long COVID symptoms and COVID-19 complications - Office for National Statistics [Internet]. 2020 Dec [cited 2021 Dec 13]. Available from:https://www.ons.gov.uk/news/statementsandletters/theprevalenceoflongcovidsymptomsandcovid19complications.

15. Sudre CH, Murray B, Varsavsky T, Graham MS, Penfold RS, Bowyer RC, et al. Attributes and predictors of long COVID. Nat Med. 2021;27(4):626–31.

16. Kumar A. Shadow of Long Covid: Why India needs to prepare for long-term effects of coronavirus - Coronavirus Outbreak News. INDIA TODAY [Internet]. 2021 Apr 28 [cited 2021 Dec 13]; Available from: https://www.indiatoday.in/coronavirus-outbreak/story/long-covid-india-corona-cases-affects-experts-infections-post-recovery-symptoms-1795987-2021-04-28.

17. MoHFW | Home [Internet]. 2021 [cited 2021 Dec 13]. Available from: https://www.mohfw.gov.in/.

18. Dwivedi S. Coronavirus: Delhi’s Post-Covid Clinic for Recovered Patients With Fresh Symptoms Opens. NDTV NEWS [Internet]. [cited 2021 Dec 13]; Available from: https://www.ndtv.com/delhi-news/coronavirus-delhis-post-covid-clinic-for-recovered-patients-with-fresh-symptoms-opens-2282756.

19. Ministry of Health and Family Welfare. National Comprehensive Guidelines for Management of PostCovid Sequelae [Internet]. 2021 [cited 2021 Dec 13]. Available from: https://www.mohfw.gov.in/pdf/NationalComprehensiveGuidelinesforManagementofPostCovidSequelae.pdf.

20. Obesity and overweight [Internet]. Who.int. 2021 [cited 18 December 2021]. Available from: https://www.who.int/news-room/fact-sheets/detail/obesity-and-overweight.

21. Global COVID-19 Clinical Platform Case Report Form (CRF) for Post COVID condition (Post COVID-19 CRF) [Internet]. 2021 Feb [cited 2021 Dec 13]. Available from: https://www.who.int/publications/i/item/global-covid-19-clinical-platform-case-report-form-(crf)-for-post-covid-conditions-(post-covid-19-crf-).

22. Michelen M, Manoharan L, Elkheir N, Cheng V, Dagens A, Hastie C, et al. Characterising long COVID: a living systematic review. BMJ Global Health. 2021;6(9):e005427.

23. Chopra N, Chowdhury M, Singh AK, Ma K, Kumar A, Ranjan P, et al. Clinical predictors of long COVID-19 and phenotypes of mild COVID-19 at a tertiary care centre in India. Drug Discov Ther. 2021;15(3):156–61.

24. Naik S, Haldar SN, Soneja M, Mundadan NG, Garg P, Mittal A, et al. Post COVID-19 sequelae: A prospective observational study from Northern India. Drug Discov Ther. 2021;15(5):254–60.

25. Fatima G, Bhatt D, Idrees J, Khalid B, Mahdi F, Mehdi F. Elucidating Post-COVID-19 manifestations in India. medRxiv [Internet]. 2021 [cited 2021 Dec 13]; Available from: https://www.medrxiv.org/content/10.1101/2021.07.06.21260115v1.

26. Budhiraja S, Aggarwal M, Wig R, Tyagi A, Mishra R, Mahajan M, et al. Long Term Health Consequences of COVID-19 in Hospitalized Patients from North India: A follow up study of upto 12 months. medRxiv [Internet]. 2021 [cited 2021 Dec 13]; Available from: https://www.medrxiv.org/content/10.1101/2021.06.21.21258543v1.

27. Mandal S, Barnett J, Brill SE, Brown JS, Denneny EK, Hare SS, et al. ‘Long-COVID’: a cross-sectional study of persisting symptoms, biomarker and imaging abnormalities following hospitalisation for COVID-19. Thorax. 2021;76(4):396–8.

28. Nehme M, Braillard O, Alcoba G, Perone SA, Courvoisier D, Chappuis F, et al. Covid-19 symptoms: Longitudinal evolution and persistence in outpatient settings. Ann Intern Med. 2021;174(5):723–5.

29. Rao GV, Gella V, Radhakrishna M, Kumar J, Chatterjee R, Kulkarni A v, et al. Post-COVID-19 symptoms are not uncommon among recovered patients-A cross-sectional online survey among the Indian population. medRxiv [Internet]. 2021 [cited 2021 Dec 13]; Available from: https://www.medrxiv.org/content/10.1101/2021.07.15.21260234v1.

30. Cabrera Martimbianco AL, Pacheco RL, Bagattini ÂM, Riera R. Frequency, signs and symptoms, and criteria adopted for long COVID-19: A systematic review. Int J Clin Pract. 2021;75(10):e14357.

31. Iqbal FM, Lam K, Sounderajah V, Clarke JM, Ashrafian H, Darzi A. Characteristics and predictors of acute and chronic post-COVID syndrome: A systematic review and meta-analysis. EClinicalMedicine. 2021;36:1–13.

32. Iwu CJ, Iwu CD, Wiysonge CS. The occurrence of long COVID: a rapid review. Pan Afr Med J 2021;38(65):1–12.

33. Anjana NK, Annie TT, Siba S, Meenu MS, Chintha S, Anish TS. Manifestations and risk factors of post COVID syndrome among COVID‐19 patients presented with minimal symptoms – A study from Kerala, India. J Family Med Prim Care 2021;10(11):4023–9.

34. Rudroff T, Kamholz J, Fietsam AC, Deters JR, Bryant AD. Post-COVID-19 Fatigue: Potential Contributing Factors. Brain Sci. 2020;10(12):1015.

35. Augustin M, Schommers P, Stecher M, Dewald F, Gieselmann L, Gruell H, et al. Post-COVID syndrome in non-hospitalised patients with COVID-19: a longitudinal prospective cohort study. Lancet Reg Health Eur. 2021;6:100122.

36. Blomberg B, Mohn KGI, Brokstad KA, Zhou F, Linchausen DW, Hansen BA, et al. Long COVID in a prospective cohort of home-isolated patients. Nat Med. 2021;27(9):1607–1613.

